# The Impact of Extracorporeal Shockwave Therapy on the Reported Pain Levels of Chronic Patients in a Clinical Setting

**DOI:** 10.1101/2020.12.06.20244996

**Authors:** Edward J. Cremata, Edward E Cremata, Ulyss Bidkaram, Bryce Brown, Allan Radman

## Abstract

**Background:** A review of literature for in-office, low to medium energy (.04mj/mm^2^ to .4mj/mm^2^) Extracorporeal Shockwave Therapy (ESWT) shows a substantial body of evidence suggesting strong efficacy and safety for the use of this form of Acoustic Compression Therapy. Much of this evidence is focused on the treatment of a specific region of the body, such as lateral epicondylitis, plantar fasciitis, and shoulder tendinopathies. This evaluation is designed to address the clinical utility of low to medium energy ESWT in an outpatient health care office setting, including delivery to multiple regions of the body, and for patients considered good candidates based on the failure of at least six months of prior conservative care.

**Methods:** Ordinary least squares (OLS) models with errors clustered at the patient level estimate the association between shockwave treatments and patient-reported pain levels. Additional models utilizing polynomial treatment indicators test for a non-linear relationship between treatment number and reported pain level.

**Results:** For the sixty-one patients represented in this analysis, the mean reduction in pain was 2.3 points on a 10 point scale, representing a 47% reduction in average reported pain levels. Results suggest that each treatment is associated with a 0.33 point reduction in reported pain levels (on a 10 point scale), controlling for patient demographics and treatment intensity. Additional models utilizing polynomial treatment indicators suggest a non-linear relationship between treatment number and reported pain level, indicating that the initial benefit of treatment is a 0.67 point reduction in pain for the first treatment, and falling slightly with each subsequent treatment. A subset of patients responded to follow up requests to ascertain reported pain levels at least three months after the final treatment. All patients were contacted, out of which 24 responded, reporting average pain levels of 2.9 out of 10, a substantial improvement from initial reported pain levels following final treatment (4.0), representing a decrease of 28%.

**Conclusion:** The results suggest the use of Acoustic Compression at these doses on properly selected cases can improve clinical outcomes for conservatively treated patients who may otherwise end up requiring more aggressive measures in the absence of ESWT. Evidence reviewed suggests that continued healing time leads to further improvement.

## Introduction

Extracorporeal Shockwave Therapy (ESWT) is used to treat a wide range of pathologies. These include enthesopathies such as Plantar Fasciitis, lateral and medial epicondylitis, as well as various tendinopathies including Knee, Achilles, and shoulder tendinopathies with or without calcium deposition. In recent years, evidence for the effectiveness of Acoustic Compression has been shown for a wide range of applications, displaying particular efficacy for tendinopathies with calcific deposition (e.g Consentino et al. 2003; Vahdatpour et al., 2012).

The mechanism inhibiting further improvement in patients with enthesopathies is likely chronic fibrosis development due to repetitive injuries, with resultant loss of flexibility, scarring, and decreased perfusion. Acoustic Compression has been shown to reduce adhesion formation and reduce calcium deposition, as well as stimulating angioneogenesis, thus reversing the cause of the patient’s chronic enthesopathies. This allows for better recovery than in-office conservative measures can often provide. Dosage is another important factor in the efficacy of ESWT. In this evaluation, the highest energy doses possible (to patient tolerance) are used, with an average of 2200 shocks administered during each treatment session (range: 1500 to 5500). The energy levels used in this analysis fall into the low to medium range of those reported in the literature, particularly when articles including ESWT administered with anesthesia are considered. All patients were treated with the WellWave extracorporeal shockwave generating unit, manufactured by Richard Wolf. For this machine, therapeutic energy levels range from 0.04 mJ/mm^2^ to 0.4 mJ/mm^2^ delivered through a focused applicator.

ESWT was offered to chronic patients who did not respond to appropriate conservative treatment for at least six months. Patients were also eligible for inclusion if they experienced recurrence of a condition after a period of temporary relief, suggesting the existence of a chronic pathology consistent with an enthesopathy, such as scarring, scar contracture, or perfusion loss at the involved region. Acute and subacute conditions responding to routine in-office care (e.g. ice, ultrasound, exercise, myofascial release techniques, orthotics for plantar fasciitis, etc.) may not be good candidates for ESWT. However, the conservative low to medium energy approach used in this study allows practitioners to consider Acoustic Compression treatment earlier for patients with suboptimal responses to routine in office treatment. The six-month waiting period commonly recommended prior to more aggressive management with high energy ESWT with anesthesia was utilized in this study, despite the use of low to medium energy treatment.

For the sixty-one patients included in this analysis, the mean reduction in pain was 2.3 points on a 10 point scale, representing a 47% reduction in average reported pain levels. Ordinary least squares estimates with standard errors clustered at the patient level suggest that each treatment is associated with a 0.33 point reduction in reported pain levels (on a 10 point scale), controlling for patient demographics and the intensity of treatment. Additional models utilizing polynomial treatment indicators suggest a non-linear relationship between treatment number and reported pain, indicating that the marginal benefit of treatment begins at a 0.67 point reduction in pain after the first treatment, falling slightly for each subsequent treatment. A subset of patients responded to follow up requests to ascertain reported pain levels at least three months after the final treatment. All patients were contacted, out of which 24 responded, reporting average pain levels of 2.9. This represents a substantial and statistically significant improvement from these patient’s reported pain levels immediately following their final treatment of 4.0 on a 10 point scale, a decrease of 28%.

Based on prior evidence, in addition to results from this in-office evaluation, acoustic compression (ESWT) should be considered in patients with chronicity, recurrence, or sub-optimal recovery from in-office enthesopathies (e.g. lateral and medial epicondylitis, shoulder tendinopathies) and various tendinopathies (e.g. of the patellar tendon, Achilles tendon, and plantar fasciitis). While many patients show immediate improvement in reported pain levels, the prognosis offered to patients should include adequate healing time of at least 3-6 months following the completion of the recommended 7-weekly session protocols.

*Variance in Treatment Efficacy* Extracorporeal Shockwave Therapy (ESWT) is used to treat a wide range of injuries and in many settings, ranging from vascular abnormalities as found in erectile dysfunction to chronic calcific enthesopathies. For example, a randomized controlled trial (RCT) found that ESWT treatment lowered heel pain compared to sham interventions by a clinically relevant amounts (Gollwitzer et al., 2007). ESWT has also been found to be more effective than transcutaneous electric nerve stimulation (TENS) in the treatment of chronic calcific tendonitis of the shoulder (Pan, 2003). Additional studies have found ESWT be effective in the treatment of tendonitis of the shoulder (Consentino et al., 2003; Mouzopoulos et al., 2007), patellar tendinopathy (Leeuwen, Zwerver, and Akker-Scheek, 2012), Achilles tendinopathies (Fridman et al., 2008), chronic proximal plantar fasciitis (Malay et al., 2006), and calcifying and non-calcifying tendinitis of the supraspinatus muscle (Haake, Rautmann, and Worth, 2001), There is also evidence that low-energy ESWT can be effective in pain reduction. In an RCT conducted to estimate the impact of low energy ESWT (3000 impulses of 0.08 mJ/mm^2^) on pain due to tennis elbow present for at least 12 months, treatment was associated with a significant reduction in pain and improvement in function compared to the control group (Rompe et. al, 1996). An RCT conducted on patients with lateral epicondylitis found that low-dose ESWT treatment without anesthesia is found to significantly lower pain for at least one year, in addition to causing improvements in functional activity scores and activity specific evaluations (Pettrone & McCall, 2005). Low to medium energy type shockwave units have also been found to effectively treat calcific tendinitis of the shoulder (Cacchio et al., 2006), calcaneal enthesophytosis (Cosentino et al., 2003; Rompe et al., 1996), plantar fasciitis (Moretti et al., 2006; Vahdatpour, 2012), and patellar tendinopathy (Furia et al., 2012).

In addition to the location and severity of injury, previous literature has identified the energy level used during treatment to be a significant factor in the efficacy of ESWT. The effects of this technology are generally dose-dependent, with higher energies yielding better results, such as less recurrence of calcification and pain (Peters, et. al, 2004). Given the importance of dose dependency, we chose to use a focused head that allows the application of higher directed energies to the targeted tissue, as opposed to a radial or linear head that provides a more diffuse energy, which may be more appropriate for myofascial treatment of myotendinopathy or trigger points within larger muscular regions.

For the chronic patients analyzed in this study we may expect delayed efficacy, especially when using lower to medium energies, since the removal of the fibrosis collected over time and with continued aggravations and flareups may be gradual with repeated treatments. These chronic patients may need additional treatment sessions beyond the standard protocol of seven treatments. In practice, only 4 patients of the total 61 were given more than seven shockwave treatments for any particular diagnosis. Exercise is also recommended for patients as early as day 1 in chronic cases, which is important because angiogenesis facilitated by acoustic compression depends on many factors, including the demand for oxygenation.

In the absence of ESWT treatment, many patients would otherwise consider surgical options. For example, 62% of respondents to a post-treatment survey sent after receiving ESWT indicated that they would have undergone “open or invasive” procedures in the absence of ESWT availability (Norris, Eickmier, & Werber, 2005). This suggests the potential importance of ESWT as an intermediate treatment option when conservative care has failed but before recommending more invasive procedures.

There is evidence for the effectiveness of ESWT in the treatment of many of the pathologies present in the patients in this analysis. These include pathologies of the shoulder, knee, various tendinopathies (e.g. Achilles, elbow), enthesopathies such as plantar fasciitis, and selected neuropathies such as a Morton’s neuroma and Carpal Tunnel Syndrome.

## Methods

This analysis seeks to determine the impact of ESWT on the reported pain levels of existing patients with chronic pathologies in a clinical practice setting. Sixty-one patients are included in this analysis, receiving a total of 389 treatments (often across multiple pathologies), for an average of 6.4 treatments each. For each patient, the highest dose possible maintaining patient comfort (with no sedation) is provided, with respect to both intensity and frequency (see Table 1 for specific dosages). The average patient receiving treatment was fifty-one years old, but substantial variation (s.d. = 18 years) suggests these results are externally valid to a broad range of ages.

**Table 1.** Demographic & Treatment Information for Patients.

|  | Mean | Std. Dev |
| --- | --- | --- |
| Age | 51 | 18 |
| Female | 38% | n/a |
| Average Frequency | 388 | 0.0 |
| Average Intensity | 14 | 4.8 |
| Head PC | 19 | 3.1 |
| Average Number of Pulses | 2176 | 795 |

Note: An intensity rating of 14 corresponds to 4.0 mJ/mm<sup>2</sup>.

Data were collected from all patients receiving ESWT during the study period. During the initial visit, data were collected on patient demographics (e.g. age and gender), and reported pain levels were collected after the first treatment. In addition, information was collected on the location of the injury, as well as the frequency, intensity, and number of pulses used during the treatment. After each subsequent visit, updated pain levels were recorded, as well as any modifications to the treatment itself (e.g. change in intensity level). Finally, at least three months after treatment was completed, each patient was contacted to provide a final pain rating, to allow for analysis of both the short and long term impact of ESWT on reported pain levels.

As can be seen in Table 2, the most commonly treated pathologies in this analysis are in the thoracic and shoulder regions, in addition to the treatment of plantar fasciitis and injuries to the Lumbo-Pelvic region. The number of ESWT treatments provided to address injuries to the bicep and Achilles tendon is insufficient to allow these regions to be analyzed separately (2 and 6 observations, respectively), but these observations are included in the overall estimate of the relationship between ESWT and pain levels.

**Table 2.**
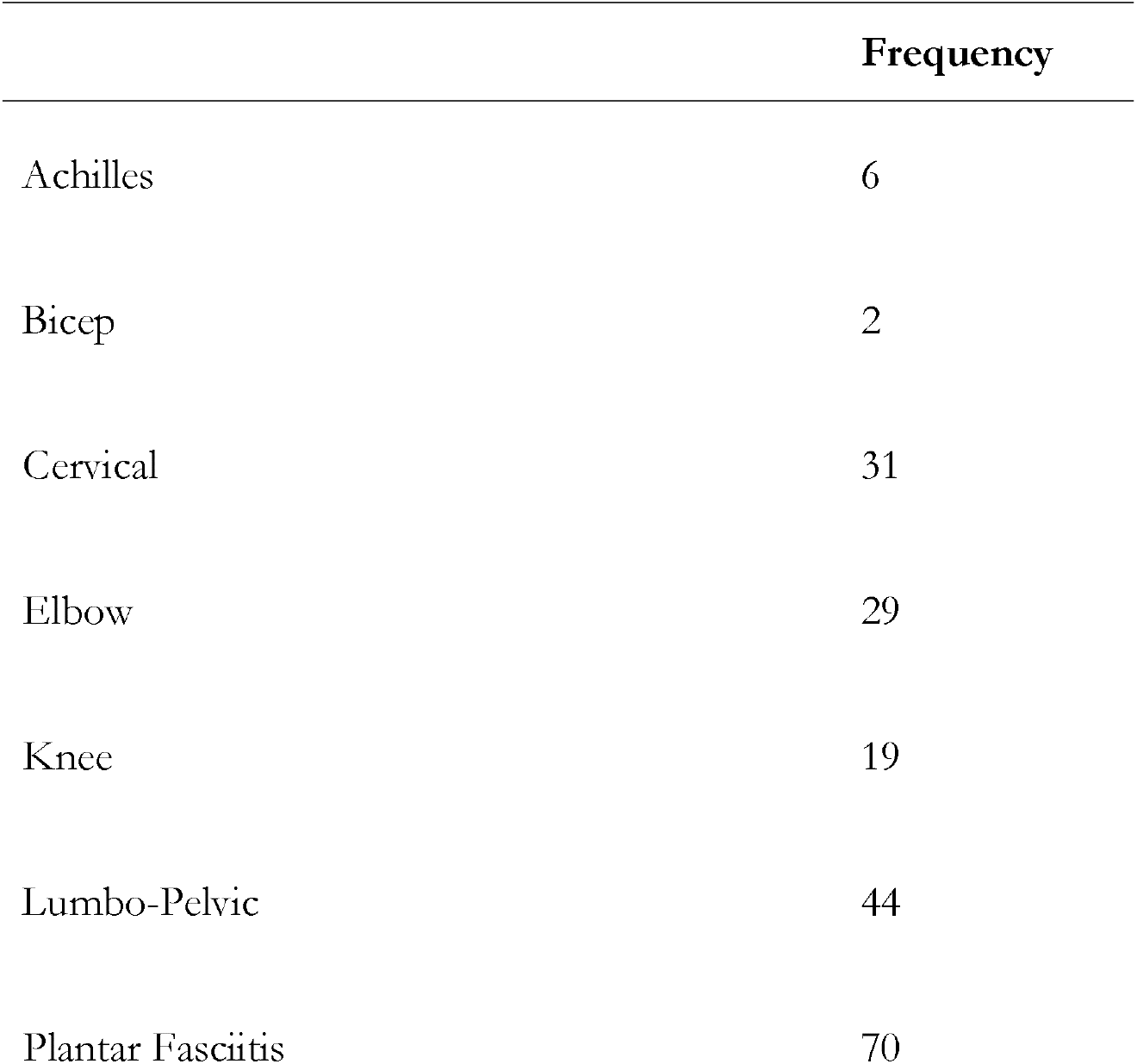
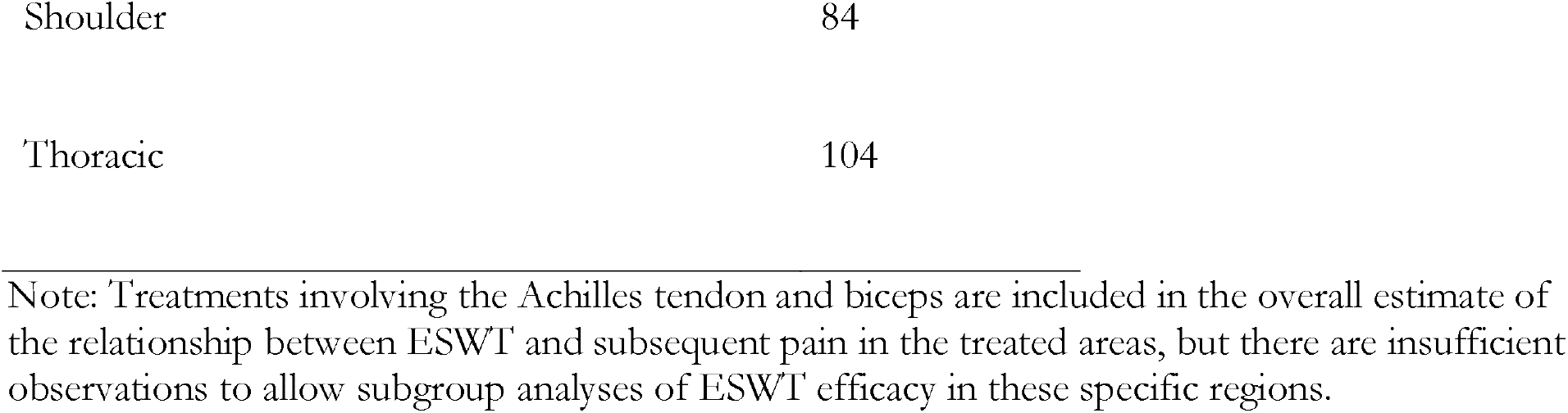
Conditions Treated by ESWT by Number of Treatments.

We see in Table 3 that a total of eighty-eight regions are treated at least once across the sixty-one patients. A large majority of patients continued to receive treatment through at least the second treatment period (93%), and over half (63%) of pathologies were addressed through at least the fourth visit. Average reported pain levels fall through the first six treatment periods, however it is important to note that the patients and injuries represented for each treatment session are not comparable. A simple comparison of average pain levels over time captures both the impact of ESWT as well as a “composition effect,” the impact of different patients and pathologies being treated for different lengths of time. For example, the rise in average reported pain levels in the seventh treatment may be due to the fact that only patients with more severe injuries were considered good candidates for acoustic compression therapy beyond the sixth treatment. In contrast, many patients stopped receiving treatments before the end of the protocol due to elimination of reported pain in the affected region.

**Table 3.** Reported Pain Levels of Patients by Number of Treatments.

| **Reported Pain**<br>**Level: 1 to 10 (n)** | **Mean** | **Std. Dev** |
| --- | --- | --- |
| All Treatments (389) | 3.9 | 2.2 |
| 1<sup>st</sup> Treatment (88) | 4.9 | 2.2 |
| 2<sup>nd</sup> Treatment (82) | 4.2 | 2.1 |
| 3<sup>rd</sup> Treatment (72) | 3.7 | 2.1 |
| 4<sup>th</sup> Treatment (55) | 3.2 | 2.2 |
| 5<sup>th</sup> Treatment (39) | 3.2 | 2.1 |
| 6<sup>th</sup> Treatment (21) | 2.8 | 1.8 |
| 7<sup>th</sup> Treatment (15) | 3.3 | 2.1 |
| 8<sup>th</sup> Treatment (7) | 2.4 | 1.9 |
| Long Term Follow Up (24) | 2.9 | 2.0 |

Note: The number of observations exceeds the number of patients due to some patients receiving ESWT treatments for multiple pathologies. In addition, 3 patients received a total of 10 treatments beyond the eighth by request. Analyses are also run excluding these patients (as their treatment exceeded the pre-determined protocol), and the results do not change.

All data preparation and analyses are conducted using the statistical software package Stata 14 (StataCorp, College Station). Given that the focus of this analysis is a “real world” evaluation of the impact of ESWT on patients in a clinical practice, randomization of patients into control and treatments groups was not possible. Despite this, there are reasons to be confident that the observed changes in reported pain levels are predominantly the result of ESWT treatment over the period of analysis. The primary justification for this assumption is that patients were selected for inclusion only after a minimum of six months of traditional care was not able to provide sufficient relief. This process of patient selection is vital to our ability to interpret changes in pain level after the administration of ESWT as the causal impact of ESWT treatment, given that the primary threat to internal validity in this analysis is the potential for natural rates of recovery in reported pain levels over time. For example, if patients with acute injuries were treated with acoustic compression therapy shortly after injury, it would be impossible to separate the causal impact of ESWT from the expected reduction in reported pain levels over time due to natural healing. However, for the patients in our analysis (those with chronic conditions unresponsive to at least 6 months of traditional care), the expected natural rate of recovery is zero or near zero. Put another way, given that each patient acts as their own control (i.e. pain levels before and after each treatment are compared for each patient to determine treatment impact), it is crucial that the counterfactual expected change in pain levels in the absence of treatment is minimal. With regard to the external validity of our results, they are applicable to clinical practices with similar patient profiles and processes for the determination of treatment. In addition to estimating the aggregate impact of ESWT on pain levels, analyses are also disaggregated by injury type where sufficient sample sizes are available, better allowing clinicians to apply the results of this analysis to their own practices, patient profiles, and injury types.

An important consideration in the analysis of ESWT is how to specify the treatment variable based on expected response rates over time. We first model the impact of acoustic compression on reported pain levels to be a linear function, with each additional dose providing a similar increment of benefit. For this estimation, the relationship between the number of ESWT treatments and reported pain levels is represented by the following equation:

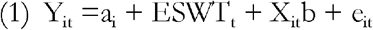

Where Y_it_ is equal to each patients reported pain level immediately after receipt of ESWT, a_i_ is the intercept, ESWT_t_ is a continuous variable equal to the number of ESWT treatments received after the first treatment, X_it_b is a vector of patient level controls (e.g. age, gender), and e_it_ represents the error term for patient i at time t (in addition to clustering standard errors at the patient level, models using robust standard errors are run and do not change the results). In the model specified above, the continuous treatment variable’s baseline is established after the first treatment (i.e. the initial “pre-treatment” pain score is taken immediately after the first ESWT treatment) to control for any short term impacts as a result of ESWT that do not indicate improvement in the underlying pathology (e.g. temporary numbness). This decision may lead the impacts reported in this paper to be a conservative estimate of the impact of ESWT, but this is necessary to ensure that our analysis is able to isolate the real, long term improvement in pain scores from any potential short-lived impact of ESWT on pain.

Additional versions of equation 1 are stratified by injury type to identify whether the treatment impact of ESWT varies by pathology. To account for the potentially non-linear relationship between the number of shockwave treatments and the outcome of interest, an extension of equation 1 is estimated including a quadratic (squared) version of the treatment variable:

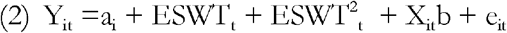

Where ESWT^2^ _t_ represents the squared continuous treatment indicator, and all other variables remain unchanged. Results for baseline and expanded models are presented in Table 4, while subgroup analyses by injury type are presented in Table 5.

**Table 4.** Impact of ESWT Treatment on Patient Reported Pain Levels.

|  | (1) Treatment Indicator Only | (2) Treatment Indicator + Demographic Controls | (3) Treatment Indicator + Demographic & Intensity Controls | (4) Non-Linear Treatment Indicator + Demographic & Intensity Controls |
| --- | --- | --- | --- | --- |
| Treatment | -0.31**<br>(0.05) | -0.34**<br>(0.05) | -0.33**<br>(0.05) | -0.67**<br>(0.15) |
| Treatment Squared |  |  |  | 0.04*<br>(0.02) |
| Patient Age |  | 0.01*<br>(0.005) | 0.01*<br>(0.005) | 0.01*<br>(0.005) |
| Patient Female |  | 1.1**<br>(0.23) | 1.0**<br>(0.26) | 1.0**<br>(0.25) |
| Treatment Intensity |  |  | -0.02<br>(0.03) | -0.02<br>(0.02) |
| Adjusted R<sup>2</sup> | 0.08 | 0.15 | 0.16 | 0.17 |
| N - Observations | 389 | 389 | 389 | 389 |

Note: Standard errors provided in parentheses below estimates. \* - significant at 5% level, \*\* - significant at 1% level.

**Table 5.**
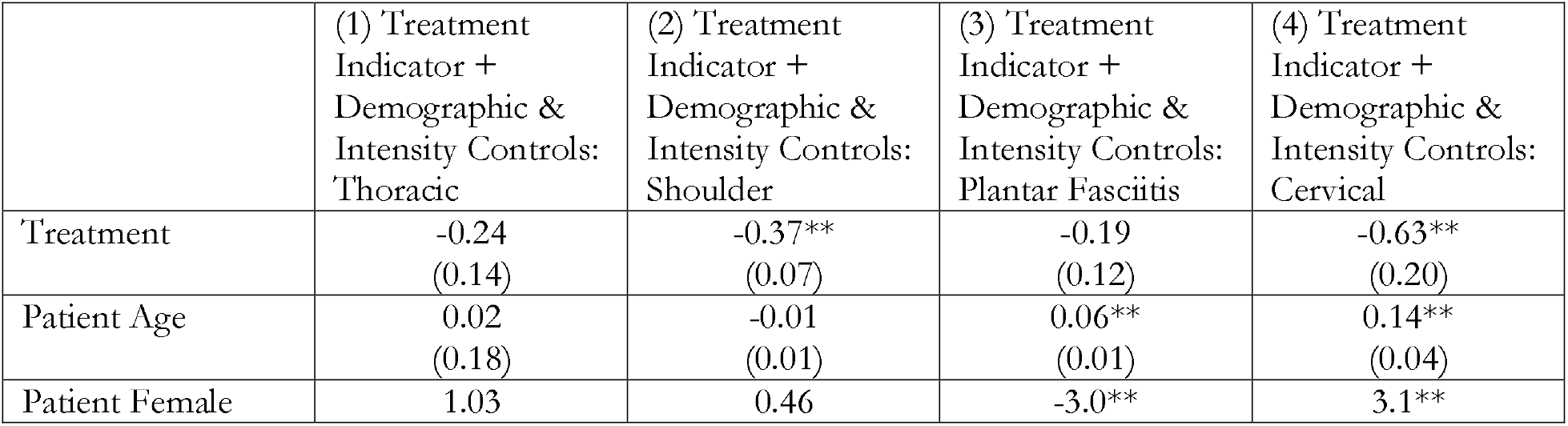
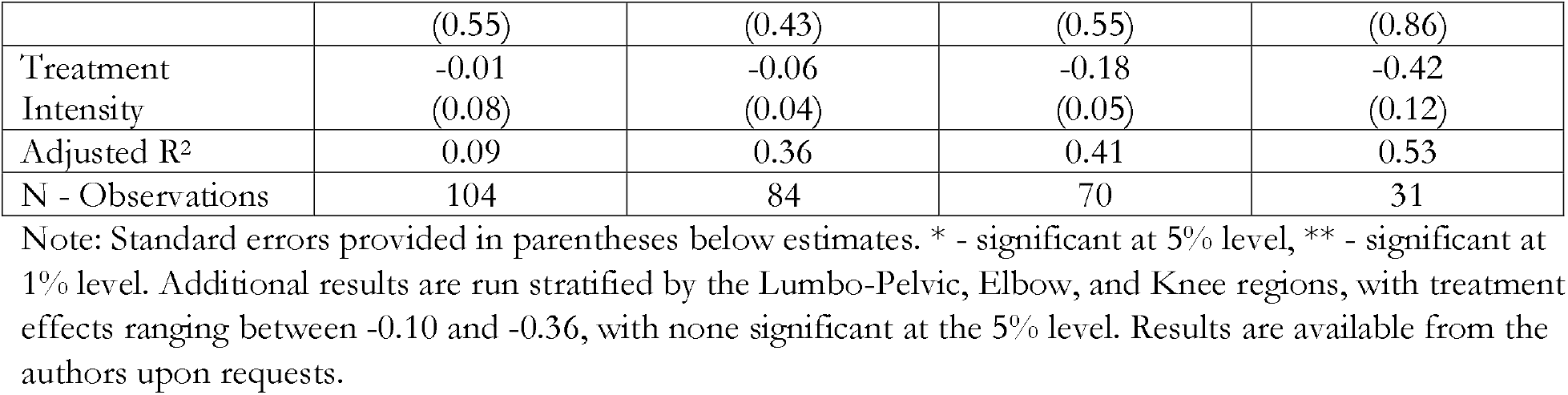
Impact of ESWT Treatment on Patient Reported Pain Levels Stratified by Region Treated.

## Results

The primary results are presented in Table 4 below. Column 1 provides an estimate of the relationship between ESWT treatment and pain levels from a bivariate regression (no controls). We see that each ESWT treatment is associated with a statistically significant 0.31 point reduction in pain. The inclusion of patient demographic controls increases the estimated impact of treatment on pain levels to 0.34 points per treatment, as seen in column 2. Controlling for the intensity level used during treatment does not substantially alter the relationship between treatment and reported pain levels (column 3). In column 4, the relationship between the number of treatments and reported pain levels is found to be significantly non-linear, with a large initial reduction in pain levels after the first treatment (−0.67 points) that falls as the number the number of treatments provided increases. This finding is consistent with established treatment guidelines to initially prescribe a limited number of acoustic compression treatments and monitor patient response. Across all models, older patients report higher levels of pain on average (0.01 points per year increase in age), females report higher pain levels (1.0 – 1.1 points), and treatment intensity has no impact on efficacy. However, it should be noted that intensity levels were provided to patient tolerance, and patient tolerance could be related to pain levels in unobserved ways. Therefore, this analysis does not suggest that the relationship between treatment intensity and ESWT impact reported in prior literature is inaccurate.

When the full model including patient demographic and treatment intensity controls is stratified by injury type, we find substantial variation in observed efficacy. In column 1 of Table 5 we see that while the estimated relationship between treatment number and reported pain levels is negative for the thoracic region, it is smaller in absolute magnitude than the aggregate estimate and not significant at traditional levels. In contrast, each additional treatment in the shoulder region is associated with a significant 0.37 point reduction in pain levels. Similar to results found for the thoracic region, as well as for the Lumbo-Pelvic and Knee subgroup analyses (not shown), the relationship between treatment and pain is negative and insignificant for these regions. Note that the loss of statistical significance for these regions is due to both the lower estimated benefit of treatment as well as the reduction in sample size that occurs when examining only injuries in a particular region. Acoustic compression is found to have the largest positive impact for injuries located in the cervical region, with each treatment found to reduce reported patient pain levels by 0.63 points. It is interesting to note that the estimated impact of ESWT on reported patient pain levels is always negative, suggesting that even in cases where sample sizes or treatment impact are small enough to provide insignificant results, there were no treatment areas in which ESWT treatment is associated with an increase in patient reported pain levels.

## Discussion

The results presented above suggest that ESWT is a safe and effective treatment for patients with various enthesopathies that had failed prior conservative management of six months or more. As noted above, the internal validity of our analysis relies on the assumption that by choosing only patients with sustained chronic pain, any observed decrease in reported pain levels coincident with the application of ESWT is not due to natural recovery. To the extent that this assumption is not credible, the causal validity of these results is threatened. For the sixty-one patients included in this analysis, the mean reduction in pain was 2.3 points on a 10 point scale, representing a 47% reduction in average reported pain levels. Baseline estimates suggest that each treatment is associated with a 0.33 point reduction in reported pain levels (on a 10 point scale), controlling for patient demographics and treatment intensity. Additional models utilizing polynomial treatment indicators suggest a non-linear relationship between treatment number and reported pain, indicating that the initial benefit of treatment is a 0.67 point reduction in pain for the first treatment, and falling slightly with each subsequent treatment. Acoustic compression therapy provided the largest benefit for patients with injuries to the shoulder and cervical regions. Importantly, a positive relationship between ESWT treatment and reported pain level was never observed, providing evidence that acoustic compression administered according to the protocols used in this analysis is both a safe and effective option for patients. A subset of patients responded to follow up requests to ascertain reported pain levels at least three months after the final treatment. All patients were contacted, out of which 24 responded, reporting average pain levels of 2.9 out of 10. This represents a substantial and statistically significant improvement from these patient’s reported pain levels following their final treatment of 4.0, representing a decrease of 28%.

## Conclusion

The results suggest that the use of Acoustic Compression at these doses on properly selected cases can improve clinical outcomes for conservatively treated patients who may otherwise end up requiring more aggressive measures in the absence of ESWT. These results, in conjunction with prior evidence on the efficacy of lower intensity acoustic compression, suggest that clinicians should consider in-office ESWT of low to medium intensity after a period of non-responsiveness to traditional conservative management.

## Data Availability

Individual patient data is HIPPAA protected and not available to be made public. Sample pain scale and patient intake documentation available upon request.

## List of Abbreviations

ESWT: Extracorporeal Shockwave Therapy

## Declarations

### Ethics approval and consent to participate

This study was approved by Life Chiropractic College West, 25001 Industrial Blvd, Hayward, CA 94545. IRB reference number PN 2012-10.

### Consent for publication

Not applicable

### Authors’ contributions

## Acknowledgements

EJC designed the study, provided all statistical analyses and interpretations of the data.

EEC set up all clinical protocols, trained all participants in ESWT protocols, and standardized functionally capacity-based pain scale interpretations to reduce subjectivity.

UB coordinated all patient care and trained all treatment providers for consistency. He established treatment protocols and provided a large portion of the treatment for patients.

BB provided treatment and maintained all treatment records in a secure, locked environment. He consulted with EEC regularly regarding protocols and data collection and provided all initial analyses for data interpretation done by EJC.

AR provided treatment and designed and assured data collection integrity and collection. He came in with a background in this area as a mechanical engineer. He participated in data collection design and acted as the liaison between the treatment staff and I (EEC) the onsite supervisor who communicated directly with EJC throughout the study.

Not applicable.

Material availability is available from the corresponding author at.

## References

1. Consentino, R. et al. Extracorporeal shock wave therapy for chronic calcific tendinitis of the shoulder: single blind study. Annals of Rheumatoid Diseases. 2003;62:248–250.

2. Vahdatpour B, Sajadieh S, Bateni V, Karami M, Sajjadieh H. Extracorporeal shock wave therapy in patients with plantar fasciitis. A randomized, placebo-controlled trial with ultrasonographic and subjective outcome assessments. Journal of Research in Medical Sciences□: The Official Journal of Isfahan University of Medical Sciences. 2012;17(9):834–838.

3. Gollwitzer, H. et al. Extracorporeal Shock Wave Therapy for Chronic Painful Heel Syndrome: A Prospective, Double Blind, Randomized Trial Assessing the Efficacy of a New Electromagnetic Shock Wave Device. The Journal of Foot and Ankle Surgery. 2007: (46), 5, 348–357.

4. Pan, P. et al. Extracorporeal Shock Wave Therapy for Chronic Calcific Tendinitis of the Shoulders: A Functional and Sonographic Study. Archives of Physical Medical Rehabilitation. 2003: vol. 84. 988–993

5. Mouzopoulos, G. et al. Extracorporeal shock wave treatment for shoulder calcific tendonitis: a systematic review. Skeletal Radiology. 2007;36:803–811.

6. Leeuwen, M.T., Zwerver, J., & Akker-Scheek. Extracorporeal shockwave therapy for patellar tendinopathy: a review of the literature. British Journal of Sports Medicine. 2009: 43;163–168.

7. Fridman, R. et al. Extracorporeal Shockwave Therapy for the Treatment of Achilles Tendinopathies: A Prospective Study. Journal of the American Podiatric Medical Association. 2008;98; 466–468.

8. Malay, D. et al. Extracorporeal Shockwave Therapy Versus Placebo for the Treatment of Chronic Proximal Plantar Fasciitis: Results of a Randomized, Placebo-Controlled, Double-Blinded, Multicenter Intervention Trial. The Journal of Foot and Ankle Surgery. 2006. 45(4); 196–210.

9. Haake, M., Rautmann, M., and Wirth T. Extracorporeal Shock Wave Therapy versus Surgical Treatment in Calcifying Tendinitis and Non Calcifying Tendinitis of the Supraspinatus Muscle. European Journal of Orthopedic Traumatology. 2001; 11: 1 –4.

10. Rompe, JD, Hopf, C., Kullmer, K., et al. Analgesic Effect of Extracorporeal Shock-wave Therapy on Chronic Tennis Elbow. Journal of Bone and Joint Surgery. 1996: 78-B:233–7.

11. Pettrone FA, McCall BR. Extracorporeal shock wave therapy without local anesthesia for chronic lateral epicondylitis. Journal of Bone and Joint Surgery Am. 2005;87-A(6):1297–1304.

12. Cacchio, A. et al. Effectiveness of Radial Shock-Wave Therapy for Calcific Tendinitis of the Shoulder: Single-Blind, Randomized Clinical Study. Physical Therapy. 2006; 86: 672–682.

13. Moretti, B et al. Extracorporeal shock wave therapy in runners with a symptomatic heel spur. Sports Medicine. 2006. 14;1029–1032.

14. Furia, J. et al. A single application of low-energy radial extracorporeal shock wave therapy is effective for the management of chronic patellar tendinopathy. Knee Surgery, Sports Traumatology, Arthroscopy. 2012.

15. Peters, J., et al. Extracorporeal shock wave therapy in calcific tendinitis of the shoulder. Skeletal Radiology. 2004: 33:712–718.

16. . Norris, M., Eickmeier, K., and Werber, B. Effectiveness of Extracorporeal Shockwave Treatment in 353 Patients with Chronic Plantar Fasciitis. Journal of the American Podiatry Medical Association 2005: 95(6): 517–524.

